# Treatment of Shengqingtongqiao Decoction for mild cognitive impairment of white matter lesions: Study protocol for a randomized, double-blind, double-dummy, parallel controlled trial

**DOI:** 10.1101/2023.08.06.23293609

**Authors:** YueChan Zhang, XueYi Han, JieQing Zhang, ZiJun Wei, JiaNing Mei, Xie Long, XiaoMin Zhen, YunYun Zhang

## Abstract

**Background:** White matter lesions(WML) is an important cause of mild cognitive impairment(MCI). Ginkgo biloba extracts (GBTs) are widely used to treat cognitive dysfunctions. But the treatment of MCI is still limited. Shenqingtongqiao Decoction(SQTQD), as a clinical empirical formula, has received good feedback in treating MCI of WML. However, there was a lack of solid clinical research on SQTQD in treating MCI. The purpose of this study is to evaluate the efficacy of SQTQD in the MCI patients of WML.

**Methods:** This is a randomized, double-blind, double-dummy, parallel-controlled trial. 80 participants will be assigned to receive SQTQD granules plus GBTs mimetics or SQTQD mimetic granules plus GBTs in a 1:1 ratio. The trial will last 24 weeks, including a 12-week intervention and 12-week follow-up. The primary outcome is MoCA and AVLT. The secondary outcome is a neuropsychological battery (including MMSE, SCWT, TMT, DST, SDMT, BNT, VFT, and CDT), quality of life(BI, ADL, and FAQ scores), emotion assessment(PHQ-9, GAD-7 score), Fazekas and ARWMCs scale, and fMRI. Researchers will record any adverse events throughout the trial.

**Discussion:** This study will provide evidence to evaluate the efficacy and safety of SQTQD for MCI of WML compared with GBTs.

**The trial is registered at Chinese Clinical Trial Registry:** Chinese Clinical Trial Registry (Number: ChiCTR2300068552)

## Introduction

White matter lesions (WML), known as white matter hyperintensities (WMH) and leukodystrophy, were presented by Hachinski in 1987^[1]^ . WML is used to describe the diffuse and symmetrical patchy or mottled cerebral white matter lesions around the ventricles and centrum semiovale, which are characterized by hyperintensities on T2-weighted or fluid-attenuated inversion recovery images^[2]^. The incidence of WML in the elderly population is 39%-96%, increasing with age^[3]^. The pathogenesis of WML remains unclear. The mian pathological features of WML are mainly pale myelin, demyelination, oligodendrocyte apoptosis, and vacuolization^[4]^. Chronic ischemia is considered the cause. Cerebral small vessel disease and cerebral amyloid angiopathy are risk factors for WML. As a subtype of cerebral small vessel disease, WML not only increases the risk of first or recurrent ischemic stroke, intracerebral hemorrhage, dementia, and mortality^[5]^, but also are progression markers ^[6][7]^ and independent predictor ^[8][9]^ of cognitive dysfunction.

Mild cognitive impairment (MCI) defines an intermediate stage between normal aging and dementia, which is a risk factor for dementia in many studie^[10]^. The pooled prevalence of MCI among Chinese elderly was 15.4% in 53 studies with 123,766 subjects^[11]^. Nowadays, there is no effective pharmacological intervention for MCI ^[12]^. However, the treatment can provide significant benefits to patients by alleviating the symptom. Potential agents for MCI treatment include AChE-I, glutamate receptor modulators, antioxidants, anti-inflammatory drugs, nootropics, immunomodulators, mainly amyloid antibodies, secretase inhibitors and Ginkgo biloba.

Ginkgo biloba tablets (GBTs), containing flavonoids, terpene lactones, and ginkgolic acids, are widely used for the treatment of cognitive dysfunctions^[13]^.The review by Carlo Tomino et al.^[14]^thought EGb 761®, one of Ginkgo biloba extracts, benefits on cognitive decline stabilization or slowing down, on ADL and neuropsychiatric deficit for MCI, Alzheimer’s and dementia patients. GBTs have been demonstrated to have antioxidative activities, which can enhance microcirculation, affect neurotransmitter levels, enhance neuroplasticit, and provide neuroprotection. Min Zhan et al.^[15]^ searched for EGb-related trials up to March 1, 2021, in four Chinese databases, three English databases, and clinical trial registry platforms. This meta-analysis showed that EGb may be an effective and safe treatment in improving MMSE, MOCA, ADL, and BI for vascular cognitive impairment patients within three months of diagnosis from. A systematic review by Cooper et al.^[16]^ found no evidence from 41 RCTs that any specific treatment was effective in reducing the transition rate from MCI to dementia. Therefore, it is essential to develop and evaluate new promising treatments for MCI.

Based on the theory of TCM, the pathogenesis of WML with MCI is deficiency of Qi and blood, and blocking of collaterals due to phlegm and blood stasis, which leads to the brain not getting enough nutrition. SQTQD is our empirical prescription for treating WML with MCI clinically, which is a compound formula composed of Dangshen 、 Huangqi 、 Danggui 、 Chuangxiong 、 Baizhi 、 Shichangpu 、 Chenpi 、 Baishao (Table 1).

**Table 1.** Standard formulation and mechanism of SQTQD.

| Pinyin name | Latin name | Doses,g | pharmacological activities |
| --- | --- | --- | --- |
| **Dangshen** | Codonopsis pilosula | 6 | reducing the tau phosphorylation via up-regulating PP2A phosphatase activity<sup>[i]</sup><br>delaying senility, regulating stomach contraction and resisting ulcer<sup>[ii]</sup> |
| **Huangqi** | Astragalus mongholicus | 6 | regulating immune function, and anti-aging, antitumor, reducing blood sugar, lowering blood lipid, anti-fibrosis, antibacterial, radiation protection, and antiviral effects<sup>[iii][iv]</sup> |
| **Danggui** | Angelica sinensis | 6 | regulating hematopoietic, antioxidant, and immunoregulatory activities<sup>[v]</sup><br>invigorating the blood, promoting circulation, lubricating the intestines, regulating menstruation as well as relieving pain, treating female irregular menstruation and amenorrhea<sup>[vi]</sup> |
| **Chuangxiong** | Ligusticum chuanxiong | 6 | having antiatherosclerosis, antimigraine, antiaging, and anticancer properties<sup>[vii][viii]</sup> |
| **Baizhi** | Angelica dahurica | 6 | having anti-inflammation, anti-tumor, anti-oxidation, analgesic activity, antiviral and anti-microbial effects, effects on the cardiovascular system, neuroprotective function, hepatoprotective activity, effects on skin diseases<sup>[ix]</sup> |
| **Shichangpu** | Acori Tatarinowii Rhizoma | 6 | having antifibrillar amyloid plaques, anti-tau phosphorylation, and anti-inflammatory effects<sup>[x][xi]</sup> |
| **Chenpi** | Citrus Reticulata | 3 | having hypolipidemic, antiplatelet activity, antioxidant and anti-inflammatory effects<sup>[xii]</sup><br>drying dampness and resolving phlegm<sup>[xiii]</sup> |
| **Baishao** | Paeoniae Radix<br>Alba | 3 | having anti-inflammation, pain easing, liver protection,<br>and anti-oxidation effects<sup>[xiv]</sup> |

The functions of SQTQD are nourishing qi and blood, and unblocking brain collaterals by removing stasis caused by phlegm and blood stasis. Previous studies have shown that the pharmacological effects of these herbs are related to cognition or its related mechanisms.

Although we have observed the effect of the compound formula clinically, no controlled study has been carried out. Therefore, we conducted this study to evaluate the impact of SQTQD on WML patients with MCI.

## Methods/Design

### 1.1 Study design

This study is a randomized, double-blind, double-dummy, controlled trial with a 1:1 allocation of SQTQD on treating MCI of WML. A flowchart of the study is shown in Fig.1. The evaluations and visits will be conducted according to the testing schedule in Table 2. After a recruitment period and obtaining the signed informed consent from all the participants, eligible participants will be randomized to an experimental group or control group, and receive treatment for 12 weeks, with a follow-up assessment at the 24^th^ week.

**Fig. 1.**
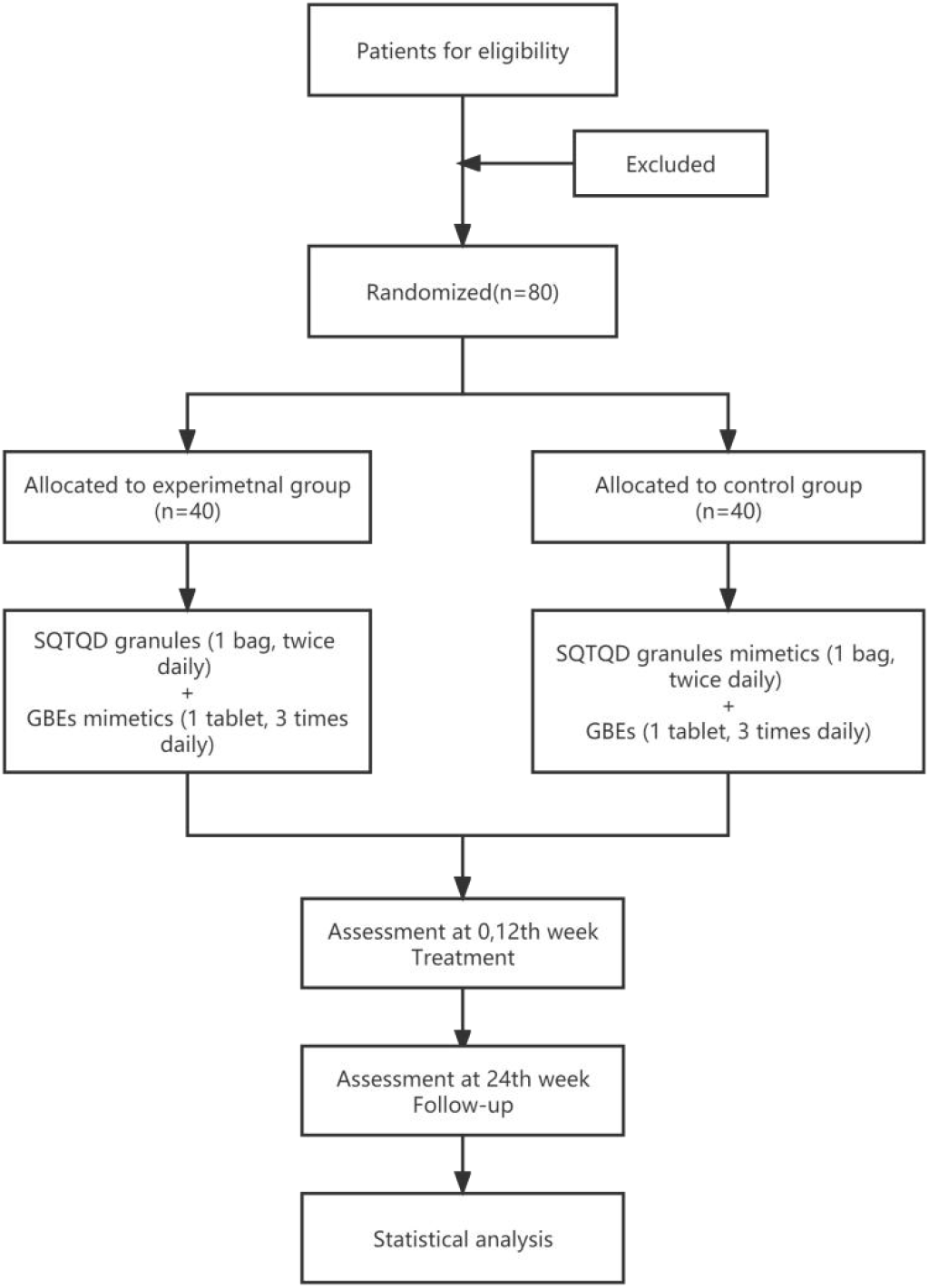
Flowchart of the study

**Table 2.** Schedule of interventions and assessments.

| Trial stage | Baseline | Mid-points |  | Endpoint | Follow-up period |
| --- | --- | --- | --- | --- | --- |
| Weeks | 0 wks | 4 wks | 8wks | 12wks | 24wks |
| **Assessments** |  |  |  |  |  |
| Informed consent | √ |  |  |  |  |
| Inclusion criteria | √ |  |  |  |  |
| Exclusion criteria | √ |  |  |  |  |
| General information | √ |  |  |  |  |
| Medical history | √ |  |  |  |  |
| Vascular Risk Factors | √ |  |  |  |  |
| Spleen-deficiency syndrome integral | √ |  |  | √ | √ |
| AIS |  |  |  |  |  |
| **Intervention** |  |  |  |  |  |
| SQTQD + GBTs mimetics |  | √ | √ | √ |  |
| SQTQD mimetics + GBTs |  | √ | √ | √ |  |
| **Primary outcomes measurements** |  |  |  |  |  |
| MoCA and AVLT | √ |  |  | √ | √ |
| **Secondary outcomes measurements** |  |  |  |  |  |
| MMSE, SCWT, ST, DST, SDMT, BNT, VFT, CDT, RCFT | √ |  |  | √ | √ |
| BI, ADL, FAQ | √ |  |  | √ | √ |
| PHQ-9、GAD-7 | √ |  |  | √ | √ |
| Fazekas、ARWMCRs<sup>[xv]</sup> | √ |  |  | √ |  |
| fMRI | √ |  |  | √ | √ |
| **Safety assessments** |  |  |  |  |  |
| Blood, urine and stool routine | √ |  |  | √ |  |
| Liver and kidney function | √ |  |  | √ |  |
| Coagulation function | √ |  |  | √ |  |
| FPG | √ |  |  | √ |  |
| Electrolyte | √ |  |  | √ |  |
| ECG | √ |  |  | √ |  |
| **Efficacy evaluation** |  |  |  | √ | √ |

### 1.2 Ethics approval and consent to participates

Ethics approval was requested and approved through the Ethics Committee of Yueyang Hospital of Integrated Traditional Chinese and Western Medicine, Shanghai University of Traditional Chinese Medicine (2021-138) . The trial was registered at the Chinese Clinical Trial Registry (Number: ChiCTR2300068552). We design this protocol in accordance with the Standard Protocol Items: Recommendation for Interventional Trials(SPIRIT). The study’s aims, procedures and possible side effects will be explained to the participants. Signed informed consent will be obtained from all the participants. No participants’ names or identifying information will be released.

### 1.3 Participants

The trial will enroll the patients meeting the diagnostic criteria for cerebral WML in the Guidelines for Neuroimage Reporting published in Lancet Neurol in 2013. The presence and severity of WML had to be agreed upon by one neurologist and one radiologist. The WML severity was assessed using the scoring system of Fazekas et al. ^[17]^. After that, we will diagnose the patients with MCI according to the Diagnostic and Statistical Manual of Mental Disorders V (DSM-V)^[18]^ and 11th revision of the international classification of diseases (ICD-11)^[19]^.

#### 1.3.1 Inclusion criteria

Patients who meet the following criteria will be enrolled in this trial.

(1) Male or female, aged from 50 to 80, adequate intellectual function to provide voluntary signed informed consent;
(2) Radiological diagnosis of WML, confirmed by brain MRI in the past 3 months, and the Fazekas’ score ≥ 2 (Table 3);
(3) Clinical diagnosis of MCI according to DSM-V and ICD-11.
(4) MoCA score of 0-13 in the illiteracy population, 0-19 in the primary school group, and 0-24 in the junior high school and above group.

**Table 3.** Rating scales of Fazeka.

| **Periventricular Hyperintensity (PVH)** |  |
| --- | --- |
| **0** | absence |
| **1** | “caps” or a pencil-thin lining |
| **2** | smooth “halo” |
| **3** | irregular PVH extending into the deep white matter |
| **Deep White-Matter Hyperintense signals (DWMH)** |  |
| **0** | absence |
| **1** | punctate foci |
| **2** | beginning confluence of foci |
| **3** | large confluent areas |

#### 1.3.2 Exclusion criteria

Enrolled patients who meet the following criteria will be excluded from the trial.

(1) A medical history of cerebral hemorrhage, cerebral infarction (infarct diameter ≥15mm), neurodegenerative diseases, brain tumors, or other central nervous system diseases.
(2) Cognitive dysfunction caused by other diseases, such as CO poisoning, hydrocephalus, immune brain injury, thyroid disease, and vitamin deficiency;
(3) Those with severe cognitive dysfunction, whose MMSE score ≤17 in the illiteracy group, ≤20 in the primary school group, ≤22 points in the secondary school group, and ≤23 points in the college group;
(4) Those who are taking pills to improve cognition in the past 3 months, such as cholinesterase inhibitors, memantine, nimodipine, and Ginkgo Biloba extract;
(5) Those with insufficiency of essential organs such as the liver, kidney, heart and lung;
(6) Those who are allergic to GBE or the ingredients of this traditional Chinese medicine;
(7) Those participating in other clinical trials in the past 3 months;
(8) Those with severe mental illness;
(9) Pregnant or lactation period women

### 1.4 Recruitment

Participants will be recruited from inpatient and outpatient in the Department of Neurology, Yueyang Hospital of Integrated Traditional Chinese and Western Medicine, Shanghai University of TCM (Shanghai, China) from Jul 21st, 2022. This trial is ongoing now. The enrollment is advertised through the hospital posters and oral promotion of hospital neurologists. All patients eligible for the trial will sign the informed consent form and get a copy with the contact telephone of the investigator. Participants can enquire questions about the trial at any time.

### 1.5 Sample size calculation

The sample size was calculated based on the sample size estimation method using the mean value of two samples in noninferiority clinical trials. The previous study shows that the change of MoCA is the primary evaluation index in similar studies, so it comes out that ur-uc =0.9,σ=1.52. Therefore, the equivalence boundary value △ was set to 0.46, with unilateral α=0.025, unilateral β=0.05, unilateral μ0.025=1.960, unilateral μ0.05=1.6449, K(the ratio of the number of cases between the experimental group and the control group)=1. Put into the formula: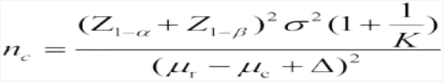.As a result, 33 participants were needed in the experimental and control groups. Considering the discontinuation of 20%, the sample size required for the study was 40 per group.

### 1.6 Randomization and blinding

A random list of numbers from 1-80 will be generated using SPSS 24.0 statistical software by an independent statistician with an allocation ratio of 1:1 for each group. Research drugs will be numerically labeled and packaged according to random numbers. The segment length was 4 to generate random numbers corresponding to the case number. According to the visit sequence, participants will be allocated randomly into experimental or control groups. The resulting random processing codes are kept in file form, with each number sealed in an envelope and kept by a person. The envelope is marked with a case number. When eligible cases entered the trial, they were treated with the same drug box according to the case number in the order of their entry.

### 1.7 Intervention

All of the participants will undergo a 12-week intervention. They will be randomly divided into 2 groups: The experimental group and Control group.

SQTQD (compositions are shown in Table 2)and its mimetics will be provided as concentrate-granules (10g/2bag) by Jiangyin Tianjiang Pharmaceutical Co., LTD (Batch number:2203303) and entrusted by the project unit (Yueyang Integrated Traditional Chinese and Western Medicine Hospital Affiliated to Shanghai University of Traditional Chinese Medicine). GBTs and GBTs mimetics are provided by Shanghai Xinyi Baluda Co., LTD. One tablet of GBTs contains 19.2mg of flavonol glucoside and 4.8mg of terpene lactone.

The participants in the experimental group will receive SQTQD granules (10g, twice daily) plus GBTs mimetics (1 tablet, 3 times daily). While in the control group, patients will receive SQTQD granules mimetics (10g, twice daily) plus GBTs (1 tablet, 3 times daily). The mimetics share the same appearance, taste, dosage, and administration methods as SQTQD granules and GBTs.

The patients will be followed up by telephone and WeChat once a month during the drug intervention period. After drug intervention, the patients will be followed up in outpatient service for the next 12 weeks.

### 1.8 Outcome evaluation

#### 1.8.1 Primary outcome

The primary outcome is the changes in cognitive function. MoCA screening tool can show a broad measure of global cognitive function. It consists of 13 tasks covering the following 8 cognitive domains, which is more sensitive and specific than MMSE in monitoring MCI^[20]^. The total score of MoCA ranges from 0 to 30, and a lower score indicates more severe cognitive impairment. AVLT, as a neuropsychological tool, tests episodic memroy to assess the cognitive function, which is mentioned in Petersen’s article^[21]^. AVLT-HuaShan version is chosen for adapting to Chinese idiom feature^[22]^.

#### 1.8.2 Secondary outcomes

The secondary outcomes as shown below:

(1) MMSE, SCWT, TMT, DST, SDMT, BNT, VFT, and CDT scale for assessing the neuropsychology and the extent of MCI;
(2) BI, ADL, and FAQ score for rating quality of life;
(3) PHQ-9, GAD-7 score for assessing emotion;
(4) Fazekas and ARWMCs^[23]^ scores for qualitatively assessing white matter hyperintensities.
(5) fMRI for assessing the changes of white matter microstructure.

#### 1.8.3 Safety assessments

The safety assessments monitoring will include the following: blood, urine and stool routine, liver and kidney function, electrolytes, coagulation function, and electrocardiogram. All side effects and adverse events will be recorded and reported to the ethics committee of Yueyang Hospital of Integrated Traditional Chinese and Western Medicine, Shanghai University of Traditional Chinese Medicine. The investigators should decide whether to stop the trial according to the severity or not.

### 1.9 Statistical analysis

All the data will be analyzed by a statistician using SPSS25.0 statistical software. For data conforming to normality and homogeneity of variance, the group t-test is used between groups, and the paired t-test is used with the group. On the contrary, Median (M), minimum (Min) and maximum (Max) were used for statistical description of data. Frequency, constituent ratio and rate were used for statistical description. All values were 2-tailed, and all analyses were significant if the value was ≤0.05.

### 1.10 Data management and quality control

To protect the anonymity and privacy of participants, participants’ names or identifying information will remain anonymous. The data management of this trial is based on an electronic data management system, and the data administrator constructs eCRF, whose content is derived from the "case report form". When all the subjects complete the study, all the case report forms are entered into the system. After they are reviewed by the inspectors and checked by the data administrator, the data are locked by the data administrator. After all the data are locked, the data administrator will import it into the designated database and submit it to the statisticians for statistical analysis.

## Discussion

MCI, an early stage of dementia, is becoming a serious public health problem in the world with the growth of the aging population^[24]^. As a common MRI finding among older adults, WML plays an important role in causing MCI^[25]^. Furthermore, many studies proved that the severity of WML is an independent predictor of decline in cognitive function and can double the risk of dementia^[26][27]^.

GBTs, which is used to treat cognitive dysfunction clinically, have been recommended to improve cognitive function and to prevent cognitive decline. Several studies have proved that GBTs have good performance in the efficacy of treating cognitive disorders, especially in ADL, neuro psychiatric deficit and memory. However, its question on efficacy to prevent cognitive decline is still open. Its poor performance on improving MCI symptoms and unknown side effects influence patients’ compliance.

TCM is an important complementary and alternative medicine used to treat MCI. SQTQD is an empirical recipe that can modify the core etiology and pathogenesis of MCI of WML according to the theory of TCM. Based on good feedback of SQTQD in clinical work, we designed this single-center trial with a small sample to determine whether SQTQD is as effective as GBTs and evaluate whether it Is worth further multicenter clinical research. To compare the effect of SQTQD on WML with MCI, we set MoCA as the primary outcome, which is more sensitive and specific in cognitive domains than MMSE. In addition, we will observe the fMRI to provide SQTQD neuromechanism in treating MCI.

The protocol still has some limitaions. The recruitment of patients is in single center with small sample. We can’t divide people into groups randomly by risk factors such as ages, genders, education. Meanwhile most outcomes will be measured by subjective scale, which may not be sensitive enough for noninferiority. Both of the limitation may affect the conclusions to some extent, causing some certain biases in the final data analysis. Although this study was designed as randomized, double-blind, double-dummy, parallel control, 24-week trial to reduce bias, the trial is still limited by its small sample. We hope that the trial will provide the evidence for the efficacy of SQTQD and GBTs in the treatment of MCI.

## Acknowledgement

This study

## Statement of Ethics

This study protocol was reviewed and approved by the Ethics Committee of Yueyang Hospital of Integrated Traditional Chinese and Western Medicine, Shanghai University of Traditional Chinese Medicine, approval number [2021–138].

## Conflict of Interest Statement

The authors have no conflicts of interest to declare.

## Funding Sources

Funded by the Science and Technology Commission of Shanghai Municipality (Grant No. 22Y11920700)

## Author Contributions

ZhangYue Chan: Conceptualization, Software, Date Curation, Investigation, Writing

XueYi Han: Conceptualization, Software, Investigation, Writing

JieQing Zhang: Conceptualization, Methodology, Writing

YunYun Zhang: Conceptualization, Writing

ZiJun Wei: Software, Investigation, Writing

JiaNing Mei: Software, Investigation, Writing

Xie Long: Conceptualization, Investigation

XiaoMin Zhen: Conceptualization, Investigation

### Data Availability Statement

The data that support the findings of this study are available on request from the corresponding author. The data are not publicly available due to privacy or ethical restrictions.

## Abbreviations

TCM =: Traditional Chinese Medicine
SQTQD =: Shengqingtongqiao Decoction
GBTs =: Ginkgo biloba tablets
WML=: White matter lesions
MCI =: Mild cognitive impairment
MRI=: Magnetic resonance imaging
MMSE=: Mini-mental State Examination
AVLT=: Auditory Verbal Learning Test
SCWT=: Stroop Color Word Test
STT=: Shape Trails Test
DST=: Digital Span Test
SDMT=: Symbol Digit Modalities Test
BNT=: Boston Naming Test
VFT=: Verbal Fluency Test
CDT=: Clock Drawing Test
RCFT=: Rey-Osterrieth complex figure test
PHQ-9=: Patient Health Questionnare
GAD-7=: Generalized Anxiety Disorder
ARWMCRs=: age-related white matter changes the rating scale
MoCA=: Montreal Cognitive Assessment
ADL=: Activity of Daily Living Scale
FAQ=: Functional Activities Questionnaire
BI=: Barthel index
fMRI=: Functional Magnetic Resonance Imaging
FPG=: fasting plasma glucose
ECG=: electrocardiogram

## Notes

### Competing Interest Statement

The authors have declared no competing interest.

### Clinical Trial

ChiCTR2300068552

### Funding Statement

This study was funded by the Science and Technology Commission of Shanghai Municipality(Grant No. 22Y11920700)

### Author Declarations

Ethics approval was requested and approved through the Ethics Committee of Yueyang Hospital of Integrated Traditional Chinese and Western Medicine, Shanghai University of Traditional Chinese Medicine (2021-138) .

